# Predictors of household food insecurity in the United States during the COVID-19 pandemic

**DOI:** 10.1101/2020.06.10.20122275

**Authors:** Brianna N. Lauren, Elisabeth R. Silver, Adam S. Faye, Jennifer A. Woo Baidal, Elissa M. Ozanne, Chin Hur

**Author notes:** Authors contributed equally (Brianna N. Lauren, Elisabeth R. Silver).

## Abstract

**Objective:** To examine associations between sociodemographic and mental health characteristics with household food insecurity as a result of the COVID-19 outbreak.

**Design:** Cross-sectional online survey analyzed using univariable tests and a multivariable logistic regression model.

**Setting:** The United States during the week of March 30, 2020.

**Participants:** Convenience sample of 1,965 American adults using Amazon’s Mechanical Turk (MTurk) platform. Participants reporting household food insecurity prior to the pandemic were excluded from analyses.

**Results:** 1,517 participants reported household food security before the COVID-19 outbreak. Among this subset, 30% reported food insecurity after the COVID-19 outbreak, 53% were women and 72% were white. On multivariable analysis, race, income, relationship status, anxiety, and depression were significantly associated with incident household food insecurity. Black respondents, Hispanic/Latino respondents, and respondents with annual income less than $100,000 were significantly more likely to experience incident household food insecurity. Individuals experiencing incident household food insecurity were 2.09 (95% CI 1.58–2.83) times more likely to screen positively for anxiety and 1.88 (95% CI 1.37–2.52) times more likely to screen positively for depression.

**Conclusions:** Food insecurity due to the COVID-19 pandemic is common, and certain populations are particularly vulnerable. There are strong associations between food insecurity and anxiety/depression. Public health interventions to increase the accessibility of healthful foods, especially for Black and Hispanic/Latino communities, are crucial to relieving the economic stress of this pandemic.

## Introduction

As the economic consequences of the COVID-19 pandemic ripple through the United States, foodbanks across the country have reported unprecedented demand, with many distribution centers falling short of community need.^(1,2)^ Unemployment has skyrocketed to Depression-era rates,^(3)^ and schools offering free or reduced-price meals have closed, potentially leaving households more vulnerable to food insecurity than ever before.

Emerging national data demonstrate an increase in household food insecurity across the country. During the week of May 7, 2020, the U.S. Census Bureau received almost 42,000 responses to their Household Pulse Survey. This survey estimated that 10% of U.S. adults experienced food scarcity in the past 7 days,^(4)^ although other national surveys have reported even higher numbers. The COVID Impact Survey, conducted by NORC at the University of Chicago for the Data Foundation, recruited a nationally representative sample of 2,190 adults during the week of April 20. Within this sample, 28% reported often or sometimes worrying about food running out within the past 30 days. In addition, 22% reported that food often or sometimes didn’t last and they didn’t have money to buy more.^(5)^ Finally, about 9,000 nonelderly adults responded to the Urban Institute’s nationally representative Health Reform Monitoring Survey between March 25 and April 10, and found that 21.9% of respondents reported household food insecurity.^(6)^

Past work examining food insecurity in the wake of major natural disasters in the US has found that risk factors for becoming food-insecure include race/ethnicity (particularly Black and Hispanic/Latino individuals), lower household income, poorer mental health, and poorer physical health.^(7,8)^ The COVID-19 pandemic has disproportionately impacted these same communities.^(9)^

The Health Reform Monitoring Survey found that Black and Hispanic adults were more than twice as likely to report household food insecurity compared to non-Hispanic white adults (33.9% and 33.3% vs 16.3%, respectively).^(6)^ As with past major disasters, the pandemic exacerbates existing systemic inequalities in access to material resources that support health and wellbeing.

Food insecurity is associated with poor health outcomes, including cardiovascular disease, body mass index over 30, and diabetes.^(10,11)^ These conditions are associated with developing severe COVID-19 symptoms.^(12)^ Thus, identifying those who are most vulnerable to household food insecurity amid the economic fallout of COVID-19 is of crucial public health importance. To this end, we conducted a cross-sectional survey of American adults to assess sociodemographic characteristics associated with incident household food insecurity in the wake of the COVID-19 outbreak.

## Methods

We conducted a cross-sectional survey of American adults through Amazon’s Mechanical Turk (MTurk), an online labor market of over 225,000 U.S. workers who complete online tasks and surveys.^(13)^ Participants were invited to take part in an online survey, administered using Qualtrics, that included questions regarding demographics, social distancing, food security before and after the COVID-19 outbreak, and anxiety and depression (described in detail below in Measures and in the Supplementary Material). Surveys were completed between March 30, 2020 – April 2, 2020, after many states had imposed stay-at-home orders.^(14)^

Participants were compensated with $0.50 for completing the survey, which took 10–15 minutes to complete.

### Measures

#### Demographics

Demographic variables included age, gender, race/ethnicity, annual household income, postal code, relationship status, employment status at time of survey, and whether participants lived with children under age 18.

#### Household Food Insecurity

We assessed household food insecurity using a validated two-item screen.^(15)^ Household food insecurity was defined as responses of “Sometimes true” or “Often true” for both items. Participants reported answers to each of these questions for the periods before and after the COVID-19 outbreak.

#### Anxiety and Depression

Anxious and depressive symptoms were assessed as part of the PROMIS-29 + 2 (PROPr) scale.^(16)^ Participant responses were expected to reflect their mental state at the time of the survey. Based on past work,^(17)^ anxiety subscale T-scores greater than or equal to 62.3 were categorized as positive for anxiety, and depression subscale T-scores greater than or equal to 59.9 were categorized as positive for depression.

#### Additional Measures

Additional survey questions asked respondents about the effect of COVID-19 on diet, exercise, and health-related quality of life. A descriptive report of findings from these measures is currently under review. These variables were not included in the present analyses as they were unrelated to our primary aims.

### Analysis

#### Univariable Analysis

The study population was restricted to respondents with household food security before the COVID-19 outbreak. We compared baseline demographic characteristics and anxiety and depression across respondents with and without incidence household food insecurity. We used Chi-square tests of independence for categorical variables, and Fisher’s exact test when expected values were less than or equal to five. Respondents with missing data for a particular analysis were excluded. Statistical significance was defined as *p* < 0.05.

#### Multivariable Analysis

We constructed a multivariable logistic regression model assessing incident household food insecurity as our outcome variable. Gender and age were included as covariables *a priori*. We included additional demographic characteristics if *p* < 0.20 on univariable analyses. Respondents with missing data for included variables were excluded from the model. Statistical testing was performed in R (Version 3.6.3).

## Results

A total of 1,965 participants across the United States completed our survey between March 30^th^ and April 2^nd^, 2020. Among the total survey respondents, 1,527 individuals (78%) reported household food security before the outbreak. Subsequent analyses were based on this subset of the population.

Within the population of food-secure participants pre-COVID-19, 44% identified as male, 72% were between the ages of 25 and 55, 72% were white, 10% were Asian, 7% were Black, and 7% were Hispanic. In addition, 48% reported an annual household income of over $50,000 and 64% reported full-time employment at the time of the survey. Roughly 30% of participants reported incident household food insecurity after the outbreak (Table 1). Figure 1 illustrates the change in household food insecurity by annual income level after the COVID-19 outbreak. Respondents at each income level reported incident household food insecurity, ranging from 19–37% of the population at different income levels.

**Table 1.**
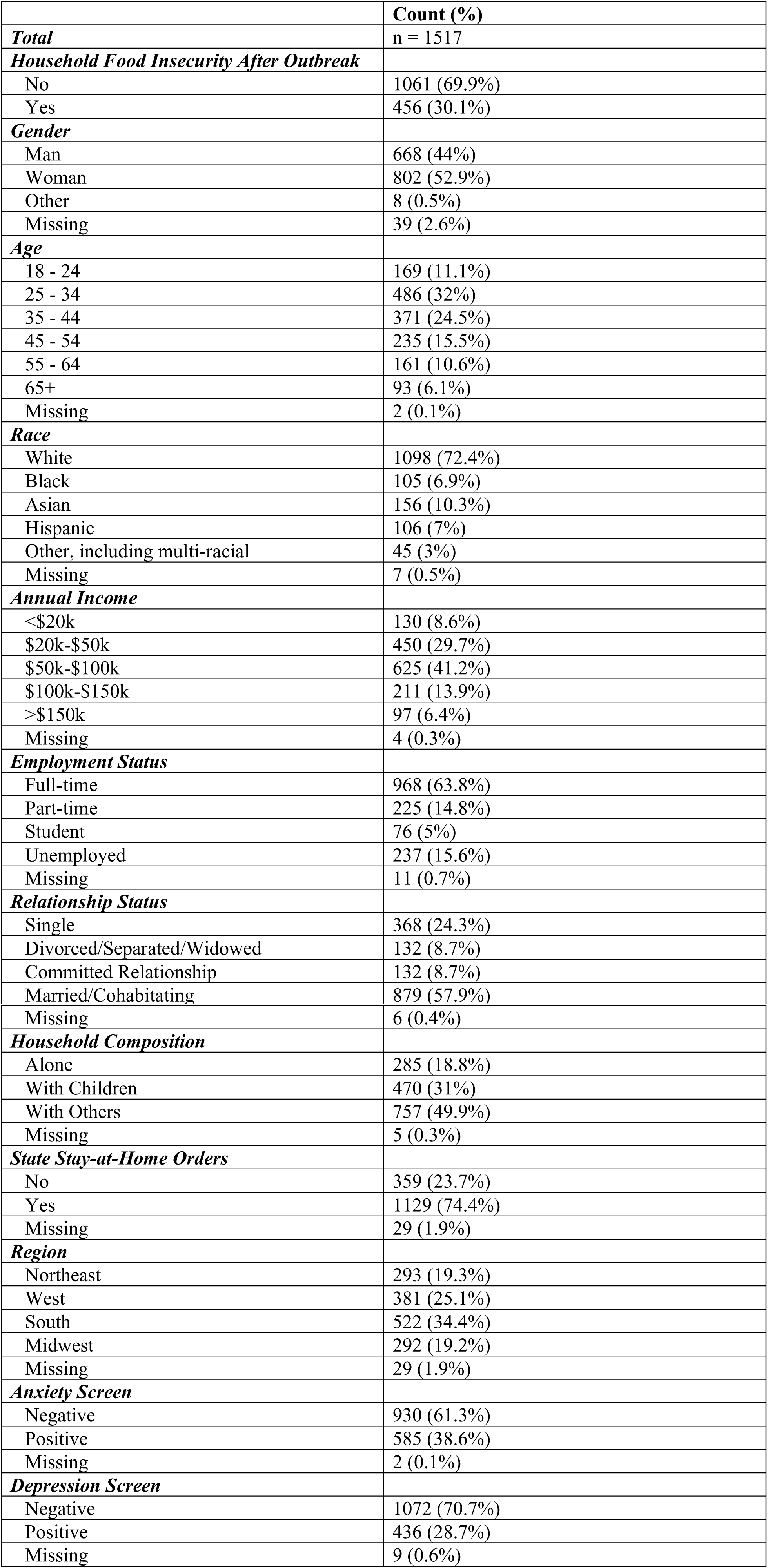
Summary of population characteristics.

|  | **Count (%)** |
| --- | --- |
| ***Total*** | n = 1517 |
| ***Household Food Insecurity After Outbreak*** |  |
| No | 1061 (69.9%) |
| Yes | 456 (30.1%) |
| ***Gender*** |  |
| Man | 668 (44%) |
| Woman | 802 (52.9%) |
| Other | 8 (0.5%) |
| Missing | 39 (2.6%) |
| ***Age*** |  |
| 18 - 24 | 169 (11.1%) |
| 25 - 34 | 486 (32%) |
| 35 - 44 | 371 (24.5%) |
| 45 - 54 | 235 (15.5%) |
| 55 - 64 | 161 (10.6%) |
| 65+ | 93 (6.1%) |
| Missing | 2 (0.1%) |
| ***Race*** |  |
| White | 1098 (72.4%) |
| Black | 105 (6.9%) |
| Asian | 156 (10.3%) |
| Hispanic | 106 (7%) |
| Other, including multi-racial | 45 (3%) |
| Missing | 7 (0.5%) |
| ***Annual Income*** |  |
| <\$20k | 130 (8.6%) |
| \$20k-\$50k | 450 (29.7%) |
| \$50k-\$100k | 625 (41.2%) |
| \$100k-\$150k | 211 (13.9%) |
| >\$150k | 97 (6.4%) |
| Missing | 4 (0.3%) |
| ***Employment Status*** |  |
| Full-time | 968 (63.8%) |
| Part-time | 225 (14.8%) |
| Student | 76 (5%) |
| Unemployed | 237 (15.6%) |
| Missing | 11 (0.7%) |
| ***Relationship Status*** |  |
| Single | 368 (24.3%) |
| Divorced/Separated/Widowed | 132 (8.7%) |
| Committed Relationship | 132 (8.7%) |
| Married/Cohabiting | 879 (57.9%) |
| Missing | 6 (0.4%) |
| ***Household Composition*** |  |
| Alone | 285 (18.8%) |
| With Children | 470 (31%) |
| With Others | 757 (49.9%) |
| Missing | 5 (0.3%) |
| ***State Stay-at-Home Orders*** |  |
| No | 359 (23.7%) |
| Yes | 1129 (74.4%) |
| Missing | 29 (1.9%) |
| ***Region*** |  |
| Northeast | 293 (19.3%) |
| West | 381 (25.1%) |
| South | 522 (34.4%) |
| Midwest | 292 (19.2%) |
| Missing | 29 (1.9%) |
| ***Anxiety Screen*** |  |
| Negative | 930 (61.3%) |
| Positive | 585 (38.6%) |
| Missing | 2 (0.1%) |
| ***Depression Screen*** |  |
| Negative | 1072 (70.7%) |
| Positive | 436 (28.7%) |
| Missing | 9 (0.6%) |

**Figure 1.**
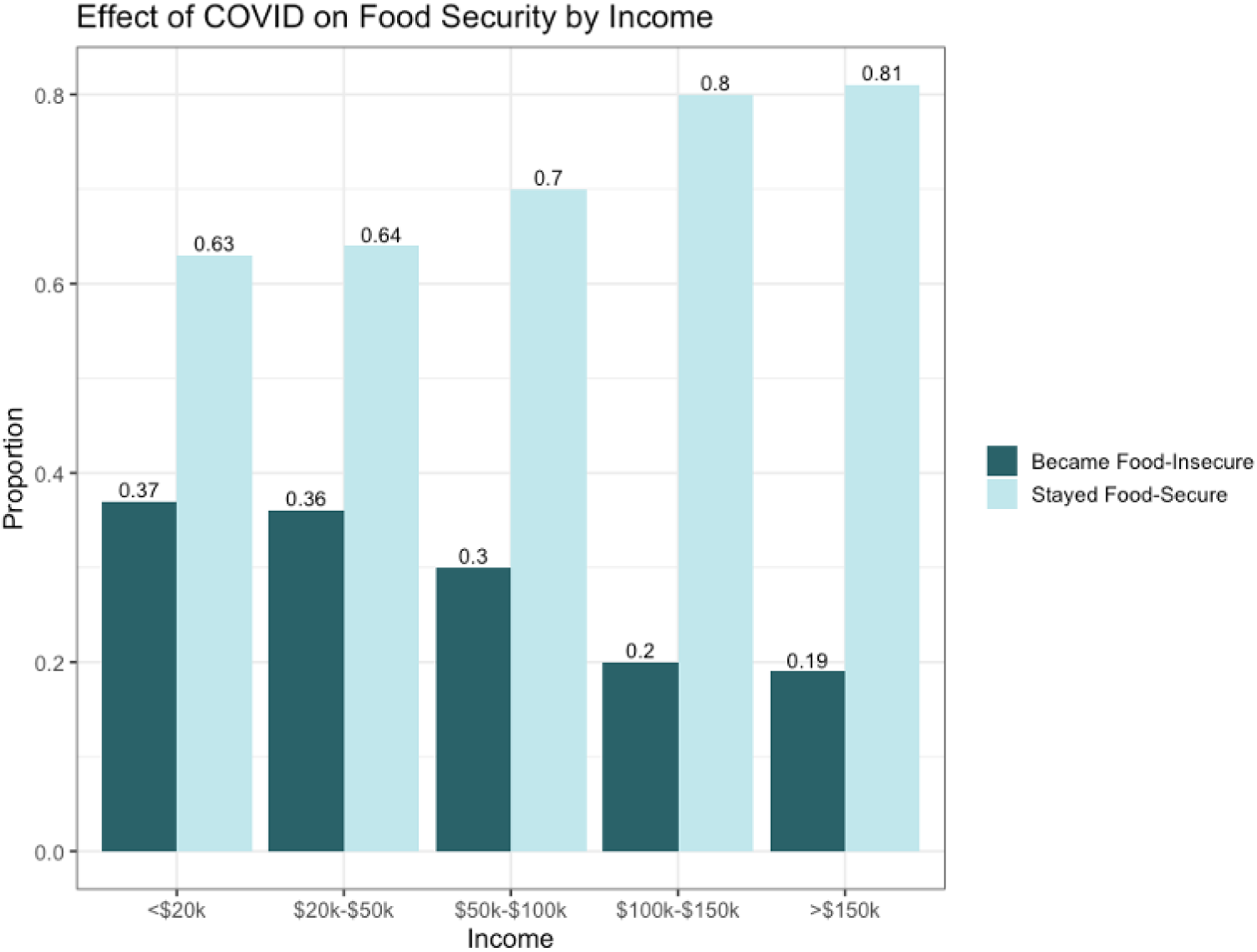
Effect of COVID on Food Security by Income.

On univariable analysis, age, race, income, anxiety, and depression were most strongly associated with incident household food insecurity (Table 2). Respondents aged 25–34 represented a larger percentage of the population with incident household food insecurity compared to the population without incident household food insecurity (38% vs 30%, respectively; *p* < 0.01). Racial/ethnic minorities and respondents with lower incomes also represented larger proportions of individuals with incident household food insecurity (*p* < 0.01). Among participants with incident household food insecurity, 57% screened positive for anxiety, compared to 31% of participants with household food security (*p* < 0.01). Similarly, 46% of participants with incident household food insecurity screened positive for depression, compared to 21.6% of participants with household food security (*p* < 0.01). Relationship status and household composition were also significantly associated with incident household food insecurity (*p* = 0.024 and *p* = 0.017, respectively).

**Table 2.** Univariable analysis results. Percentages may not sum to 100 due to missing cases.

| Characteristic |  | Became Food-Insecure<br>N (%) | Remained Food-Secure<br>N (%) | p-value |
| --- | --- | --- | --- | --- |
|  |  | 456 | 1061 |  |
| Age, years | 18 - 24 | 60 (13.2) | 109 (10.3) | <0.001 |
|  | 25 - 34 | 172 (37.7) | 314 (29.7) |  |
|  | 35 - 44 | 103 (22.6) | 268 (25.3) |  |
|  | 45 - 54 | 74 (16.2) | 161 (15.2) |  |
|  | 55 - 64 | 32 (7.0) | 129 (12.2) |  |
|  | 65+ | 15 (3.3) | 78 (7.4) |  |
| Gender | Man | 203 (45.6) | 465 (45.0) | 0.960 |
|  | Woman | 240 (53.9) | 562 (54.4) |  |
|  | Other | 2 (0.4) | 6 (0.6) |  |
| Race/Ethnicity | White | 291 (64.1) | 807 (76.4) | <0.001 |
|  | Black | 49 (10.8) | 56 (5.3) |  |
|  | Asian | 51 (11.2) | 105 (9.9) |  |
|  | Hispanic | 45 (9.9) | 61 (5.8) |  |
|  | Other, including multiracial | 18 (4.0) | 27 (2.6) |  |
| Household Income | <\$20k | 48 (10.5) | 82 (7.8) | <0.001 |
| | \$20k-\$50k | 161 (35.4) | 289 (27.3) | |
| | \$50k-\$100k | 186 (40.9) | 439 (41.5) | |
| | \$100k-\$150k | 42 (9.2) | 169 (16.0) | |
| | >\$150k | 18 (4.0) | 79 (7.5) | |
| Region | Northeast | 86 (19.3) | 207 (19.8) | 0.077 |
|  | West | 122 (27.4) | 259 (24.8) |  |
|  | South | 167 (37.5) | 355 (34.0) |  |
|  | Midwest | 70 (15.7) | 222 (21.3) |  |
| Relationship Status | Single | 96 (21.1) | 272 (25.8) | 0.024 |
|  | Divorced/Separated/<br>Widowed | 30 (6.6) | 102 (9.7) |  |
|  | Committed Relationship | 46 (10.1) | 86 (8.1) |  |
|  | Married/<br>Cohabiting | 283 (62.2) | 596 (56.4) |  |
| Employment Status | Full-time | 310 (68.3) | 658 (62.5) | 0.130 |
|  | Part-time | 65 (14.3) | 160 (15.2) |  |
|  | Student | 21 (4.6) | 55 (5.2) |  |
|  | Unemployed | 58 (12.8) | 179 (17.0) |  |
| Impact of COVID-19 on Employment | Became Unemployed | 27 (5.9) | 64 (6.1) | 1.00 |
|  | No Change in Employment | 427 (94.1) | 988 (93.9) |  |
| State Stay-at-Home Orders | Enacted | 344 (77.3) | 785 (75.3) | 0.438 |
|  | Not Enacted | 101 (22.7) | 258 (24.7) |  |
| Household Composition | Alone | 75 (16.5) | 210 (19.8) | 0.017 |
|  | With Children | 164 (36.1) | 306 (28.9) |  |
|  | With Others | 215 (47.4) | 542 (51.2) |  |
| Anxiety Screen | Positive | 259 (56.8) | 326 (30.8) | <0.001 |
|  | Negative | 197 (43.2) | 733 (69.2) |  |
| Depression Screen | Positive | 208 (45.8) | 228 (21.6) | <0.001 |
|  | Negative | 246 (54.2) | 826 (78.4) |  |

On multivariable analysis, race, income, relationship status, anxiety, and depression were all independently associated with incident food insecurity (Figure 2). Both Black and Hispanic respondents were significantly more likely to experience incident food insecurity compared to white respondents (adj OR 1.79, 95% CI 1.13–2.82 and 1.82, 95% CI 1.15–2.88, respectively). Relative to those with annual income greater than $150,000, those with annual incomes less than $100,000 were significantly more likely to experience incident household food insecurity, with the greatest effect observed for those with annual incomes less than $20,000 (adj OR 2.76, 95% CI 1.38 – 5.67). In addition, respondents who were married or cohabitating were significantly more likely to experience incident household food insecurity compared to single/casually dating respondents, with an odds ratio of 1.66 (95%CI 1.15–2.43). Finally, respondents screening positive for anxiety or depression were significantly more likely to report incident household food insecurity, with odds ratios of 2.11 (95% CI 1.58–2.83) and 1.86 (95% CI 1.37–2.52), respectively.

**Figure 2.**
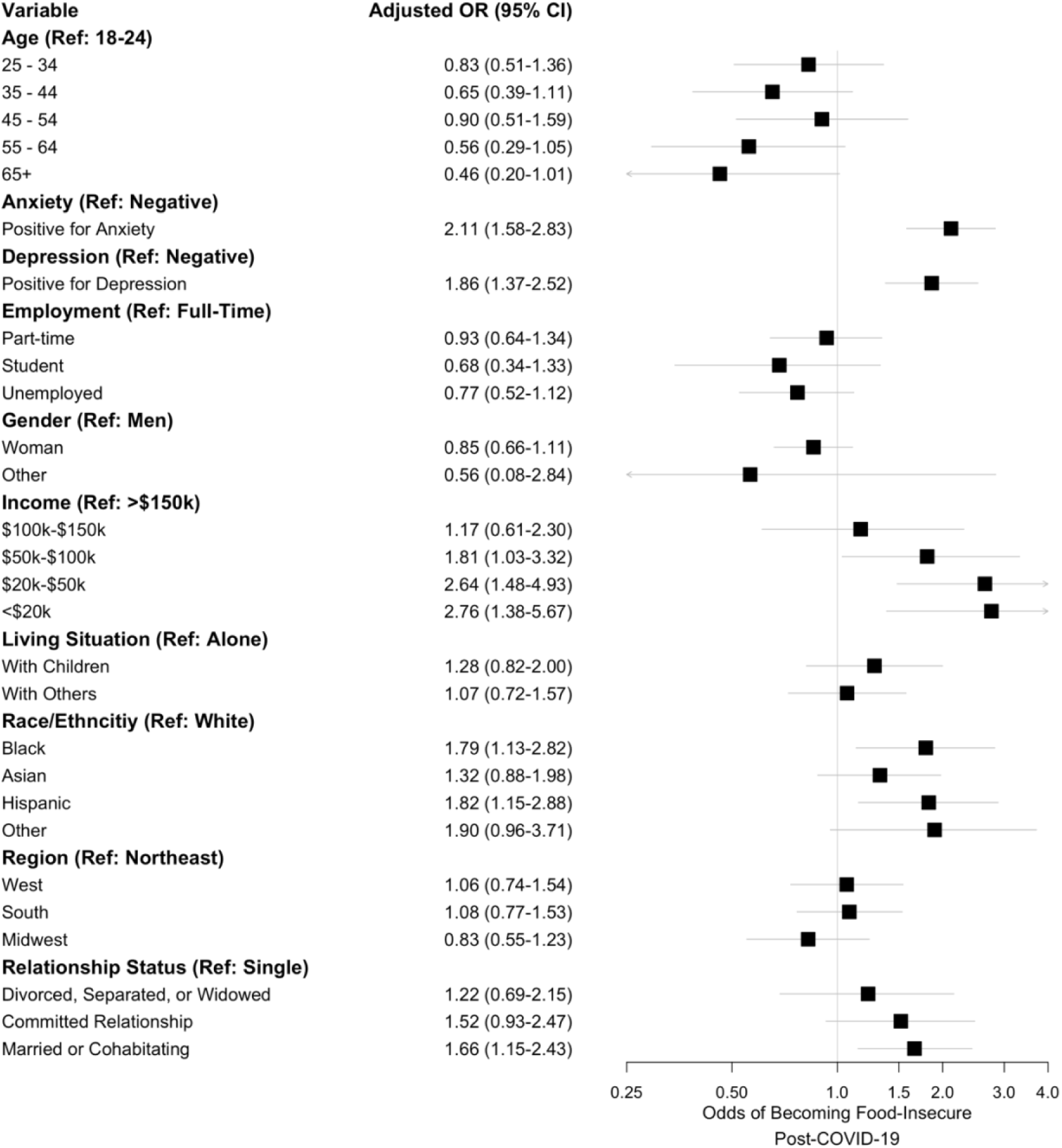
Multivariable logistic regression analysis results.

## Discussion

In this cross-sectional survey of American adults, we found that 30% of respondents became food-insecure following the COVID-19 outbreak. By contrast, the USDA reported that only 11% of total U.S. households were food-insecure in 2018.^(18)^ Risk for incident household food insecurity was significantly higher among Black and Hispanic participants, as well as participants who were married or cohabitating, or reported household incomes below $100,000 at the time of the survey. In addition, there was a significant association between incident household food insecurity and screening positive for anxiety or depression. These results suggest that the COVID-19 pandemic exacerbates existing societal inequalities, with potential downstream consequences for health and pandemic recovery efforts.

Emerging data suggest that Black and Hispanic individuals are disproportionately impacted by the COVID-19 pandemic.^(19)^ In New York City, the epicenter of COVID-19 in the US, Black and Hispanic/Latino COVID-19 patients are twice as likely to die of the disease than whites.^(9)^ Moreover, Black and Hispanic populations are disproportionately affected by cardiovascular disease,^(20)^ which is associated both with food insecurity(10,11) and severe COVID-19 disease.^(12)^ The results of our study provide further evidence of these disparities. Black and Hispanic individuals were almost twice as likely to experience incident household food insecurity, even after adjusting for other factors such as income and employment status. Targeted efforts to increase household food security among Black and Hispanic communities are needed to alleviate the disproportionate impact of COVID-19 on people of color.

In addition to highlighting existing disparities, our results underscore the far-reaching impacts of COVID-19. Among respondents with an annual household income between $50,000 and $100,000, 30% experienced incident household food insecurity after the outbreak. This proportion was lower for respondents with an annual household income above $150,000, but still remained at a surprising 19%. By contrast, the USDA reported only 5.4% of households with annual incomes at or above 185% of the federal poverty line ($47,110 for a family of four) experienced household food insecurity in 2018.^(18)^ This discrepancy highlights the extent of pandemic’s effects – many Americans are becoming newly vulnerable to household food insecurity.

Our finding regarding the relationship between food insecurity and anxiety/depression is consistent with past work documenting a bidirectional relationship between food insecurity and poor mental health and emotional wellbeing.^(21)^ More specifically, our study found that respondents experiencing incident household food insecurity were roughly twice as likely to experience anxiety or depression, even after controlling for other factors. Given that the relationship between poor mental health and food insecurity is exacerbated by social isolation,^(22)^ increasing the accessibility of mental healthcare during this time may augment more direct efforts to bolster food security.

Our study has limitations. Firstly, our cross-sectional approach precludes causality inferences, and relies on retrospective reports of food insecurity prior to the COVID-19 outbreak. Secondly, we collected data through Amazon’s Mechanical Turk, potentially creating a response bias and limiting the generalizability of our findings to the larger US population. Finally, our data were collected in late March, when the economic impacts of the outbreak were just beginning to affect many nationwide. We hypothesize that this may partially explain the unexpected result that unemployment was not a predictor of incident food insecurity. Although the validated food insecurity screening questions that we used in our study specifically referenced an ability to afford food (rather than an inability to buy food due to shortages), our finding of increased risk of food insecurity among those with annual income between $50,000-$100,000 is also surprising. Because we did not specify a time frame for these earnings, participants’ responses may reflect historical income rather than concurrent changes in annual income resulting from the pandemic. Continued research on the effects of the pandemic on food insecurity, particularly in larger community-based samples, is necessary to fully understand its impacts.

Our study quantifies the effects of the COVID-19 pandemic on household food insecurity in the US and identifies associated risk factors. Our results suggest that the economic fallout of the pandemic has severely worsened food insecurity, with minorities, low-income individuals, and those with poorer mental health at significantly higher risk. As such, targeted relief efforts are needed as we look towards economic recovery.

## Data Availability

Data are available on request to the corresponding author (Chin Hur).

## Financial Support

This research received no specific grant from any funding agency, commercial or not-for-profit sectors.

## Conflict of Interest

No personal or financial relationships to disclose (all authors).

## Authorship

BNL: Conceptualization, study design, analysis, manuscript writing and editing; ERS: conceptualization, study design, analysis, manuscript writing and editing; ASF: Analysis, manuscript editing; JAWB: Study design, manuscript editing; EMO: Study design, manuscript editing; CH: Conceptualization, study design, analysis, manuscript writing and editing, supervision.

## Ethical Standards Disclosure

This study was conducted according to the guidelines laid down in the Declaration of Helsinki and all procedures involving research study participants were approved by the Columbia University Institutional Review Board. Electronic informed agreement to participate was obtained from participants.

## Notes

### Competing Interest Statement

The authors have declared no competing interest.

### Funding Statement

No external funding was received for this research.

### Author Declarations

Columbia University Medical Center IRB (protocol #AAAS9665)

